# Inferred duration of infectious period of SARS-CoV-2: rapid scoping review and analysis of available evidence for asymptomatic and symptomatic COVID-19 cases

**DOI:** 10.1101/2020.04.25.20079889

**Authors:** Andrew W. Byrne, David McEvoy, Áine B. Collins, Kevin Hunt, Miriam Casey, Ann Barber, Francis Butler, John Griffin, Elizabeth A. Lane, Conor McAloon, Kirsty O’Brien, Patrick Wall, Kieran A. Walsh, Simon J. More

**Affiliations:** One-Health Scientific Support Unit, DAFM, Government of Ireland, Kildare Street, Dublin 2, Ireland; School of Public Health, Physiotherapy, and Sports Science, University College Dublin, Belfield, Dublin 4, Ireland; Centre for Veterinary Epidemiology and Risk Analysis, School of Veterinary Medicine, University College Dublin, Belfield, Dublin 4, Ireland; School of Biosystems and Food Engineering, University College Dublin, Belfield, Dublin 4, Ireland; School of Veterinary Medicine, University College Dublin, Belfield, Dublin 4, Ireland; Department of Agriculture, Food and the Marine, Government of Ireland, Kildare Street, Dublin 2, Ireland; Health Information and Quality Authority (HIQA), Unit 1301, City Gate, Cork, Ireland

## Abstract

**Objectives:** Our objective was to review the literature on the inferred duration of the infectious period of COVID-19, caused by SARS-COV-2 virus, and provide an overview of the variation depending on the methodological approach.

**Design:** Rapid scoping review. Literature review with fixed search terms, up to 1^st^ April 2020. Central tendency and variation of the parameter estimates for infectious period in (a) asymptomatic (b) symptomatic cases from (*i*) virological studies (repeated testing), (*ii*) tracing studies (*iii)* modelling studies were gathered. Narrative review of viral dynamics.

**Information sources:** Search strategies developed and the following searched: PubMed, Google Scholar, MedRxiv, BioRxiv. Additionally, the Health Information Quality Authority (Ireland) viral load synthesis was utilised, which screened literature from PubMed, Embase, ScienceDirect, NHS evidence, Cochrane, medRxiv and bioRxiv, HRB open databases.

**Results:** There was substantial variation in the estimates, and how infectious period was inferred. One study provided approximate median infectious period for asymptomatic cases of 6.5-9.5 days. Median pre-symptomatic infectious period across studies varied over <1-4 days. Estimated mean time from symptom onset to two negative RT-PCR tests was 13.4 days (95%CI: 10.9-15.8), but was shorter when studies included children or less severe cases. Estimated mean duration from symptom onset to hospital discharge or death (potential maximal infectious period) was 18.1 days (95%CI: 15.1–21.0); time to discharge was on average 4 days shorter than time-to-death. Viral dynamic data and model infectious parameters were often shorter than repeated diagnostic data.

**Conclusions:** There are limitations of inferring infectiousness from repeated diagnosis, viral loads, and viral replication data alone, and also potential patient recall bias relevant to estimating exposure and symptom onset times. Despite this, available data provides a preliminary evidence base to inform models of central tendency for key parameters, and variation for exploring parameter space and sensitivity analysis. Some current models may be underestimating infectious period.

## Introduction

Severe acute respiratory syndrome coronavirus 2 (SARS-CoV-2), a new coronavirus, emerged in China in late 2019.[1,2] The virus causes COVID-19, a disease characterized by variable, mainly respiratory, symptoms across cohorts, from asymptomatic cases through to mild (for example, dry cough, fever) and severe cases (for example, pneumonia).[3,4] The severity of symptoms, and their clinical outcome, have been reported to vary by age-class and whether patients have underlying comorbidities. The case-fatality rate increases with age, and are highest for those above 70 years.[5,6] There are several cases of asymptomatic test-positive patients reported in the emerging literature (e.g. [4,7,8]). Furthermore, asymptomatic (and pre-symptomatic) cases have been shown to be infectious, and secondary cases have been reported.[9,10] However, the duration of this infectious period is difficult to measure accurately, and the time course of the natural history of infection generally must be inferred indirectly, via contact tracing of cases, serial repeated diagnostic virological studies, and/or through modelling approaches. Symptomatic cases can experience an infectious pre-symptomatic period before the onset of symptoms, therefore understanding the whole infectious period for this cohort requires estimating the duration of both periods. It is essential to rapidly gain insight into this key variable impacting our understanding of COVID-19 epidemiology. Anderson et al. [11] point out one of the “key unknowns” is the infectious period for COVID-19, which they suggest may be 10 days but subject to great uncertainty.

Here we gathered data from published research from peer-reviewed and preprints from 1^st^ December to 1^st^ April 2020, to characterize the variation in the infectious duration inferred from the three lines of evidence. We also provide a narrative review of the viral dynamic literature. Our focus was on duration, relative infectiousness has been dealt with elsewhere [12,13]

The aim of this review was to provide an overview and critical appraisal of published and preprint articles and reports that assess or quantify the inferred duration of the infectious period in order to best parameterise COVID-19 epidemiological transmission models.

## Materials and Methods

### Conceptual model of population infection dynamics

Infectious period was contextualised in relation to a working conceptual model of COVID-19 disease dynamics (Figure S1). From this conceptual model, three parameters were identified as important in context of this study:

> T2, defined as: Duration of the total infectious period for asymptomatic cases, post-latent to recovery [‘recover’ in this context relates to clearing of infection]
>
> T3, defined as: Duration of pre-symptomatic infectious period for those infected individuals who subsequently develop symptoms (that is, post-latent to onset of symptoms)
>
> T5, defined as: Duration from onset of symptoms to recovery* or death.

* recovery was inferred as either the first of two clear RT-PCR tests, or hospital discharge after admission from COVID-19 related symptoms.

“Asymptomatic” case definition was interpreted pragmatically following Davies et al. [14,15], and may include very mild symptoms that may occur but are unnoticed.

T2, T3, T5 represent readily measurable parameters, but may be upper limits of infectious period, as patients may be non-infectious for a period before recovery or death. We also review evidence where infectiousness is inferred from viral shedding and contract tracing [transmission], see below.

### Literature search

A survey of the literature between 1^st^ December 2019 and 1^st^ April 2020 for all countries was implemented using the following search strategy. Publications on the electronic databases PubMed, Google Scholar, MedRxiv and BioRxiv were searched with the following keywords: “Novel coronavirus” OR “SARS-CoV-2” OR “2019-nCoV” OR “COVID-19” AND “infectious”. Additionally, national and international government reports were monitored. No restrictions on language or publication status were imposed so long as an English abstract was available. Articles were evaluated for data relating to the aim of this review; all relevant publications were considered for possible inclusion. Bibliographies within these publications were also searched for additional resources.

Manual searches of the literature was undertaken using daily updated COVID19 collections from the National Centre for Biotechnology Information (NCBI) and MedRxiv servers (https://connect.medrxiv.org/relate/content/181), respectively, searching specifically for papers relating to “infectious period” or “infectious duration” from both empirical and modelling studies.

Finally, we utilised the complementary work undertaken by the Health Information and Quality Authority (HIQA) of Ireland, specifically the evidence summaries relating to asymptomatic transmission and viral load [16,17]. The protocol for the evidence synthesis is published on the HIQA website [18]. Briefly, the evidence synthesis process included searching databases from 30^th^

December 2019 to 27^th^ March 2020 (PubMed, Embase, ScienceDirect, NHS evidence, Cochrane, medRxiv and bioRxiv, HRB open), screening, data extraction, critical appraisal and summarizing the evidence.

Our aim was to have as great a breadth for an evidential base as possible, to clarify what evidence was available to inform on the infectious period of COVID19, and to identify key characteristics of the data sources and their interpretation. Therefore, our approach is a scoping review (following [19]). However, due to the emergent nature of COVID-19, this work is considered a rapid review.[20] This paper follows the Preferred Reporting Items for Systematic Reviews and Meta-Analyses— Extension for Scoping Reviews (PRISMA-ScR) checklist.

Inclusion criteria were for papers that provided data to inform duration of infectious period based on: time from symptoms to recovery; time from symptoms to death; time from symptoms to diagnostic test clearance [≥two clear tests, defined as at least two consecutive negative reverse transcriptase polymerase chain reaction (RT-PCR) tests conducted 24 hours apart]; pre-symptomatic infectious period; time from first diagnostic test to diagnostic test clearance [≥two clear tests] for pre-symptomatic/asymptomatic cases. Inclusion criteria for viral dynamics, were papers which reported viral load via cycle threshold (Ct) values from RT-PCR testing over repeated sampling of infected patients, and studies that additional reported viral isolation.

For quality control, studies were (*i*) selected and screened initially by three members of the team from search terms outlined above (*ÁBC, KH, FB*), with parameters identified and recorded. (*ii*) This was reviewed and supplemented by manual search by a different two team members (*AWB, DM*), again with parameters identified and recorded. (*iii*) Finally, the review was then internally reviewed by an additional two members of the team (*CMc, MC*), and cross-referenced with other parameter synthesis documents being worked on by the group (*all authors*).

### Parameter comparison

#### Parameters of interest

1. *A-priori* it was decided to harvest parameter estimates for *(i)* asymptomatic, and *(ii)* symptomatic cases. As the period of infectiousness can only be estimated indirectly, parameter estimates from the literature was gathered from three different methodological approaches:Virological studies tracking patients overtime undertaking serial testing, where infectious period was inferred from diagnostic testing history and/or by virus isolation.
2. Contact tracing studies where infectiousness is inferred by infector-infectee histories and/or clusters of infection.
3. Model parameters entered into mathematical models [priors] representing explicitly infectious periods, or model parameters estimated from mathematical models [posterior estimates] estimating explicitly infectious periods

#### Visual and quantitative comparisons

To compare parameters visually, simulated distributions were estimated from the central tendencies and variation metrics described in the primary literature. To simulate data, 10,000 random variates were drawn from random number functions in Stata (ME, version 15.1; StataCorp. 2017. Stata Statistical Software: Release 15. College Station, TX: StataCorp LLC) [rnormal, rgamma]. Where possible, the distribution reported within the primary literature was used to represent the distribution (e.g. Gaussian, Gamma). Where distributional data could not be inferred, point estimates were presented.

There were adequate comparable data gathered on the duration of T5 (duration from onset of symptoms to death or recovery) from virological studies to employ a meta-analytic model. Many of the studies report different central tendency estimates, including mean and median. Variation across this central tendencies included standard deviation, range, inter-quartile range. To allow for meta-analytic comparison, the mean and standard deviations were derived for each parameter given the available central tendency and variation reported in studies based on the formulae given in Wan et al. [21].

To obtain the standard deviations from 95%CI, the method outlined in the Cochrane handbook [22] was used:

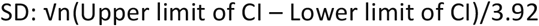

Standard Error (SE) was calculated from Standard Deviation (SD) and sample size (n), using:

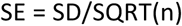

Comparisons were made using the METAAN package in Stata 15, using the random-effects (DerSimonian-Laird) model.[23] This model assumes heterogeneity between the studies; that is, it assumes that the true effect can be different for each study. The model assumes that the individual-study true effects are distributed with a variance τ^2^ around an overall true effect, but the model makes no assumptions about the form of the distribution of either the within-study or the between-studies effects. Weightings were derived from the standard error [precision] around the estimate. Comparisons were presented as forest plots. Heterogeneity between studies was tested using Cochrane’s Q; the magnitude of the heterogeneity was categorised using *i*^*2*^ as high (>75%), moderate (50-75%), or low (<50%).[24]

Variation in duration across T5 virological studies was compared using a random effects meta-regression model, using the METAREG command in Stata 15.1. The hypothesis that heterogeneity may be related to the inclusion of children or depending on symptom severity within the sample, was tested in separate univariate models. Severity was dichotomised (0/1) into studies that included patients described as having ‘mild’ or ‘mild-moderate’ symptoms, versus studies that included patients with ‘moderate-severe’ or ‘severe’ symptoms. Similarly, studies were categorised into having some samples from “children” (as reported in the paper), or wholly adult samples. These variables were then fitted as a dichotomous dummy predictor [independent] variables. The parameter estimates from the regression model was solved using restricted maximum likelihood (REML); additionally, p-values were estimated using a Monte Carlo model with 1000 permutation test.[25]

Raw patient-level data were available from three studies in relation to time from onset to hospital discharge or death (potentially inferring maximal T5 duration). To estimate the predicted mean and 95%CI duration across these studies, data were analysed using a Gaussian random effects model (using XTREG command, Stata 15), with study categories fitted as the RE. A linear regression model with ‘study’ fitted as a categorical dummy variable was used to estimate the difference between duration across study datasets. Code and data are provided in supplementary material.

### Viral dynamics

A narrative comparison of reported viral dynamics from studies that undertook serial viral load estimates from patients over their period of observation was undertaken. Trends in the literature, strength and weaknesses were identified, and a conceptual model illustrated.

## Results

### Parameter comparison

Overall, 65 parameter estimates were harvested from 48 papers (Tables 1, 2, 3).

**Table 1:**
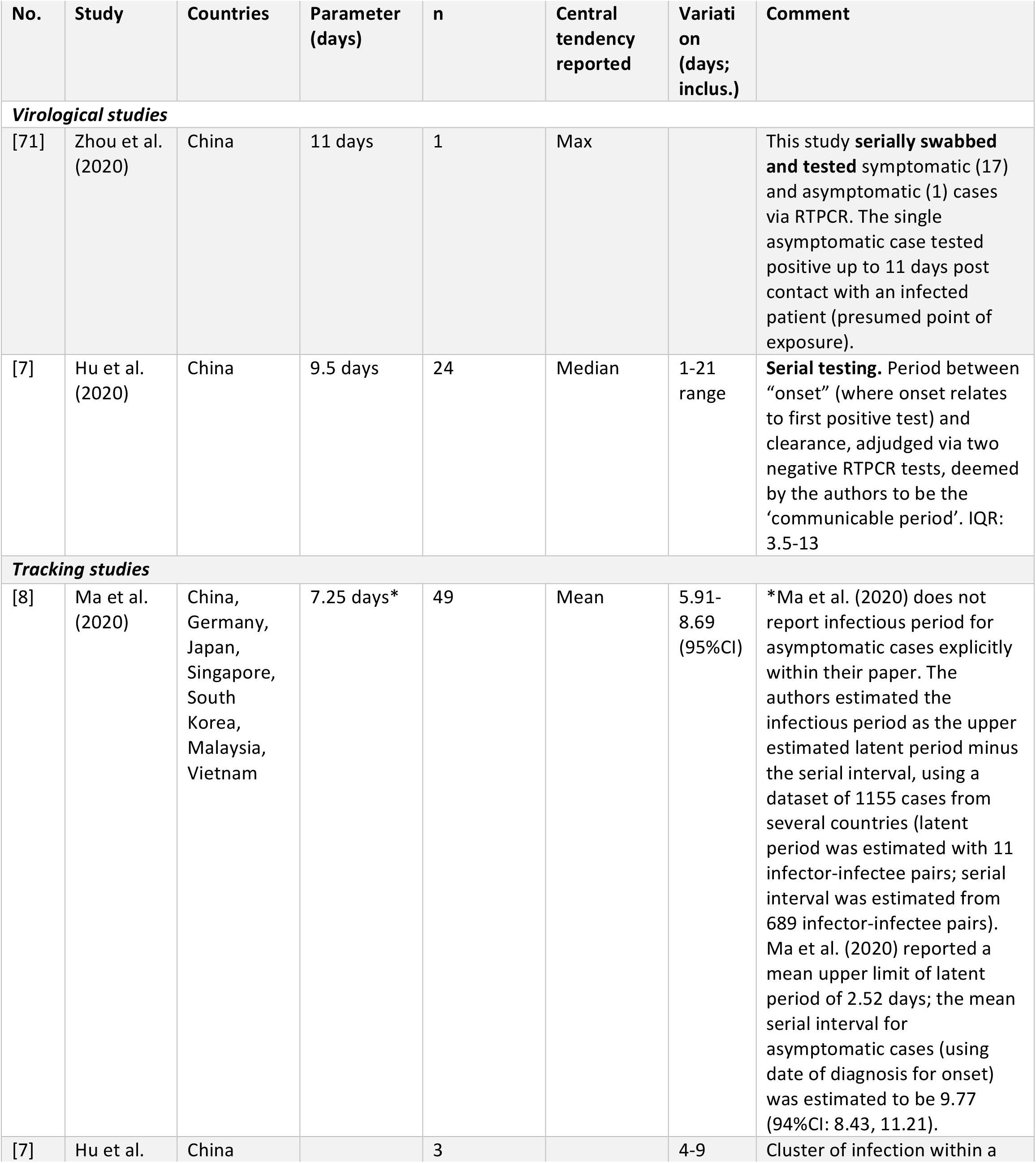
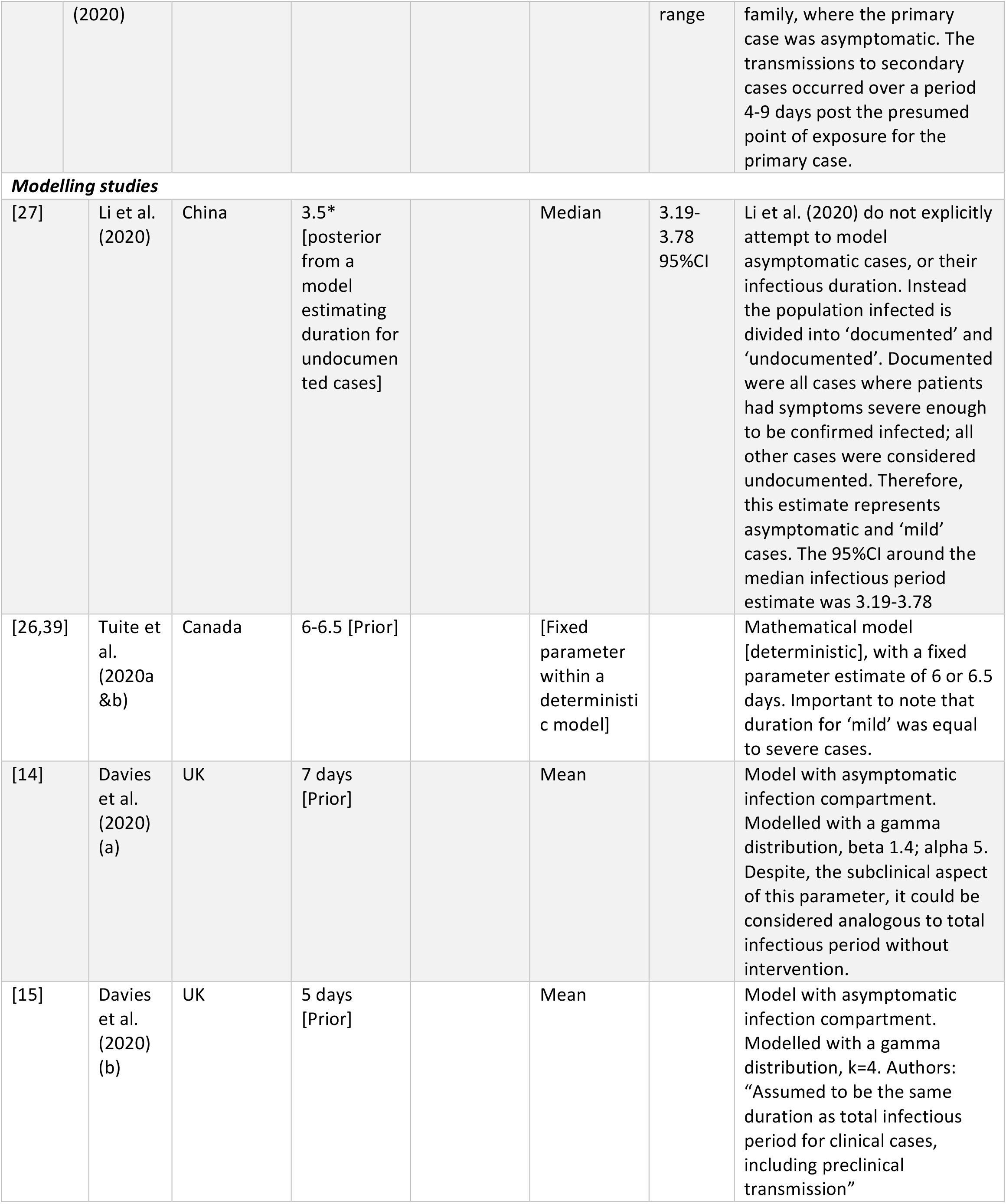
Reported infectious period (IP) for asymptomatic cases (T5 parameter) from virological studies where serial diagnostic tests were undertaken to infer IP; tracking studies where IP is inferred from contact tracing; modelling studies where IP is reported as a prior (assumed parameter value) or an posterior estimate.

**Table 2:** Reported infectious period (IP) for pre-symptomatic cases (T3 parameter) from virological studies where serial diagnostic tests were undertaken to infer IP; tracking studies where IP is inferred from contact tracing; modelling studies where IP is reported as a prior (assumed parameter value) or an posterior estimate.

|  | Study | Location | Parameter (days) | Central tendency reported | Variation (days; inclus.) | Comment |
| --- | --- | --- | --- | --- | --- | --- |
|  | ***Virological studies*** |  |  |  |  |  |
| [3] | Pan et al. (2020) | Beijing, China | 1 | Median |  | Case study of two individuals tracked due to exposure to an infected patient was **serially tested** prior to onset of symptoms. |
| [28] | Hoehl et al. (2020) | Flight from Wuhan to Germany | 1 | Median |  | Case study of **serially tested** at risk cohort flying from Wuhan to Germany. Two patients were asymptomatic test positive; additionally virus isolation was achieved, indicating potential infectiousness. |
|  | ***Tracking studies*** |  |  |  |  |  |
| [4] | Huang et al. (2020) | Nanjing, China | 4 | Median | 3-5 range | Follow-up **tracing** case study cluster of infection within a family demonstrating pre-symptomatic infection (n=10) |
| [9] | Rothe et al. (2020) | Germany | 2 | Median | 1-3 range | **Tracing** case study of a cluster of infections whereby pre-symptomatic transmission occurred (n=3). |
| [29] | He et al. (2020) | Vietnam, Malaysia, Japan, China, Taiwan, USA, Singapore | 2.3 | Mean | 95% CI, 0.8–3.0 | **Tracing** paper infector-infectee pairs. Estimated from serial interval and incubation periods. N=77 |
| [30] | Wei et al. (2020) | Singapore | 2.5 | Median | 2-3 (IQR) | **Tracing** study investigating pre-symptomatic infections from primary cases to secondary cases in 7 clusters. N=8 primary cases. T3 estimated as the min. days between transmission period (TP) and primary case symptom onset, when TP straddled >1 day. Range: 2-6 days. |
| **Modelling studies** |  |  |  |  |  |  |
| [32] | Peak et al. (2020) | Massachusetts | 0.8 [estimate] | Mean | -0.29-1.98 95% CI* | **Modelling paper** estimated under two scenarios – a serial interval of 4.8 days or 7.5 days. Under scenario one, the model estimated a period of pre-symptomatic transmission (median: 0.71). * the lower range was fixed at zero as the model allowed for no pre-symptomatic infectious case. |
| [37] | Zhu et al. (2020) | Wuhan, China | 1.0 [estimate] | Mean |  | **Modelling paper.** Model estimated point value – This is a model derived value |
| [14] | Davies et al. (2020) (a) | UK | 2.4 [prior] | Mean |  | **Modelling paper.** Gamma distribution; k=5. |
| [15] | Davies et al. (2020) (b) | UK | 1.5 [prior] | Mean |  | **Modelling paper.** Gamma distribution: k=4 |
| [26,39] | Tuite et al. (2020a & b) | Canada | 0.5, 1 [prior] | Fixed |  | **Modelling paper.** Fixed parameter within a deterministic model. |
| [72] | Ferguson et al. (2020) | UK | 0.5 [prior] | Fixed |  | **Modelling paper.** Fixed parameter within a this model, whereby infectiousness was assumed to begin 12 hours become symptoms. |
| [31] | Tindale et al. (2020) | Tianjin, China, and Singapore | 2.9-2.6 [estimate] | Mean | 1.2-8.2 mean range, depending on early or late cases, or whether in Tianjin, Singapore | Statistical **modelling** study estimating period pre-symptomatic transmission inferred from estimates of serial interval and incubation periods for populations in Tianjin and Singapore (n=228). |

**Table 3:**
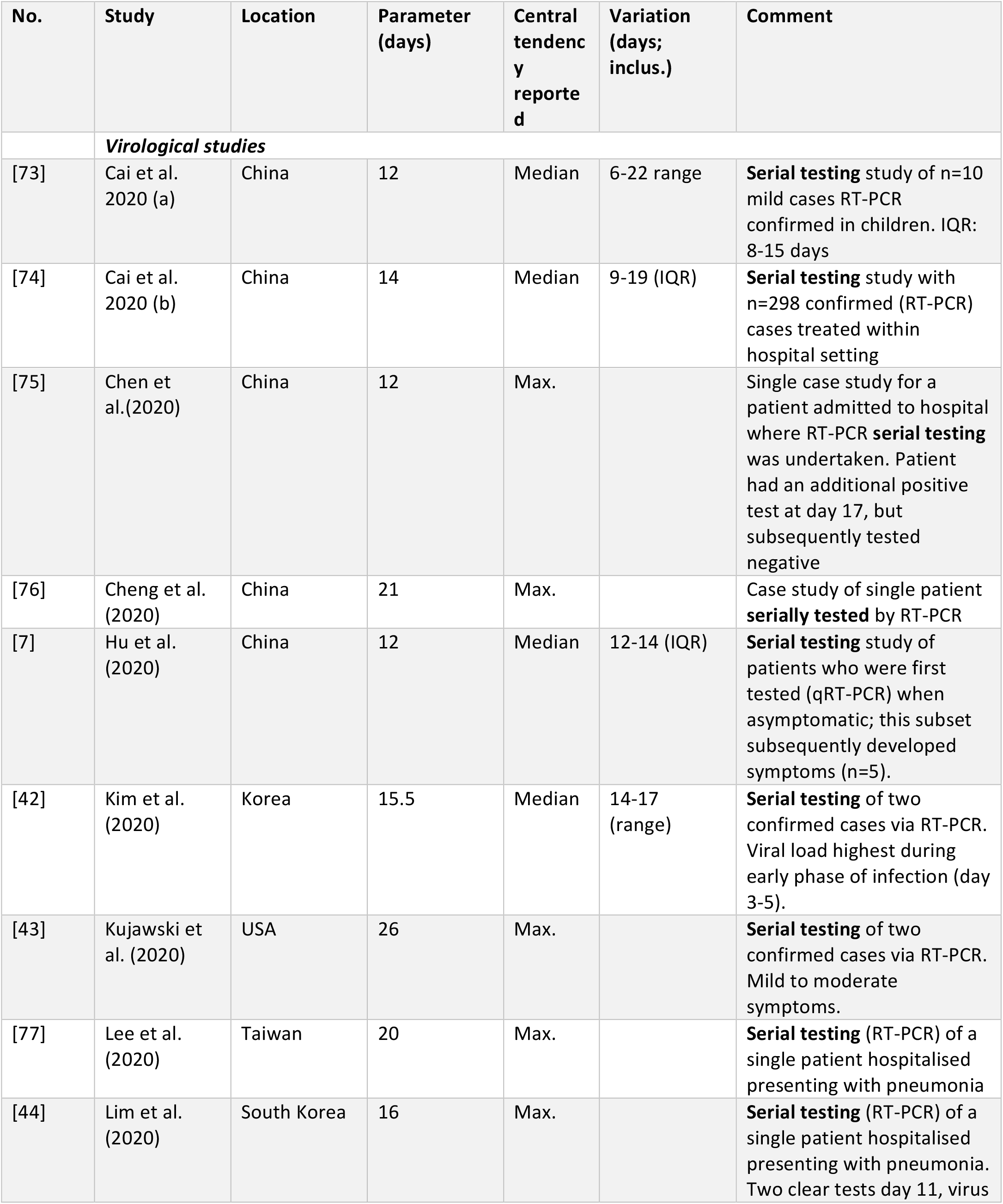
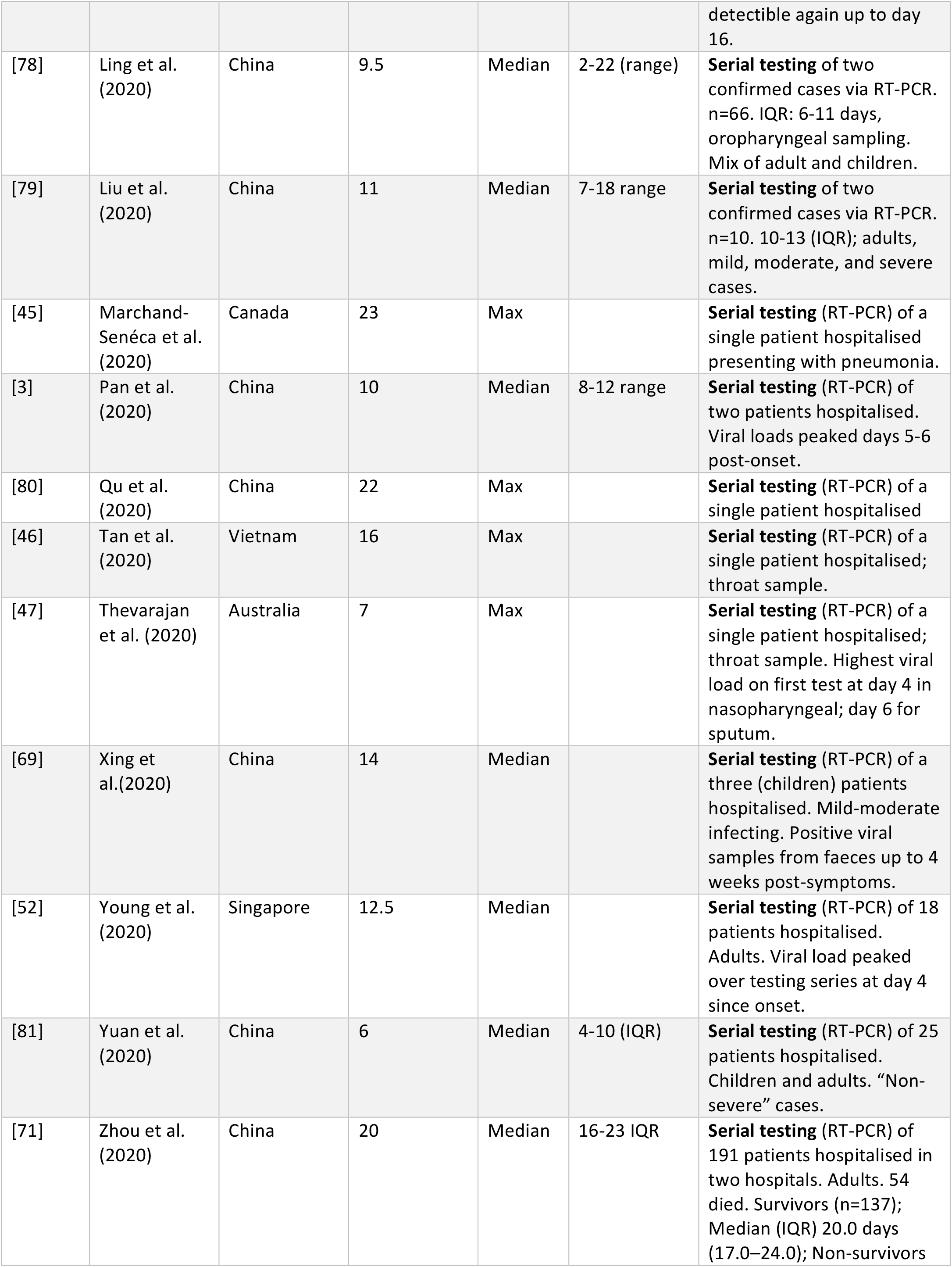
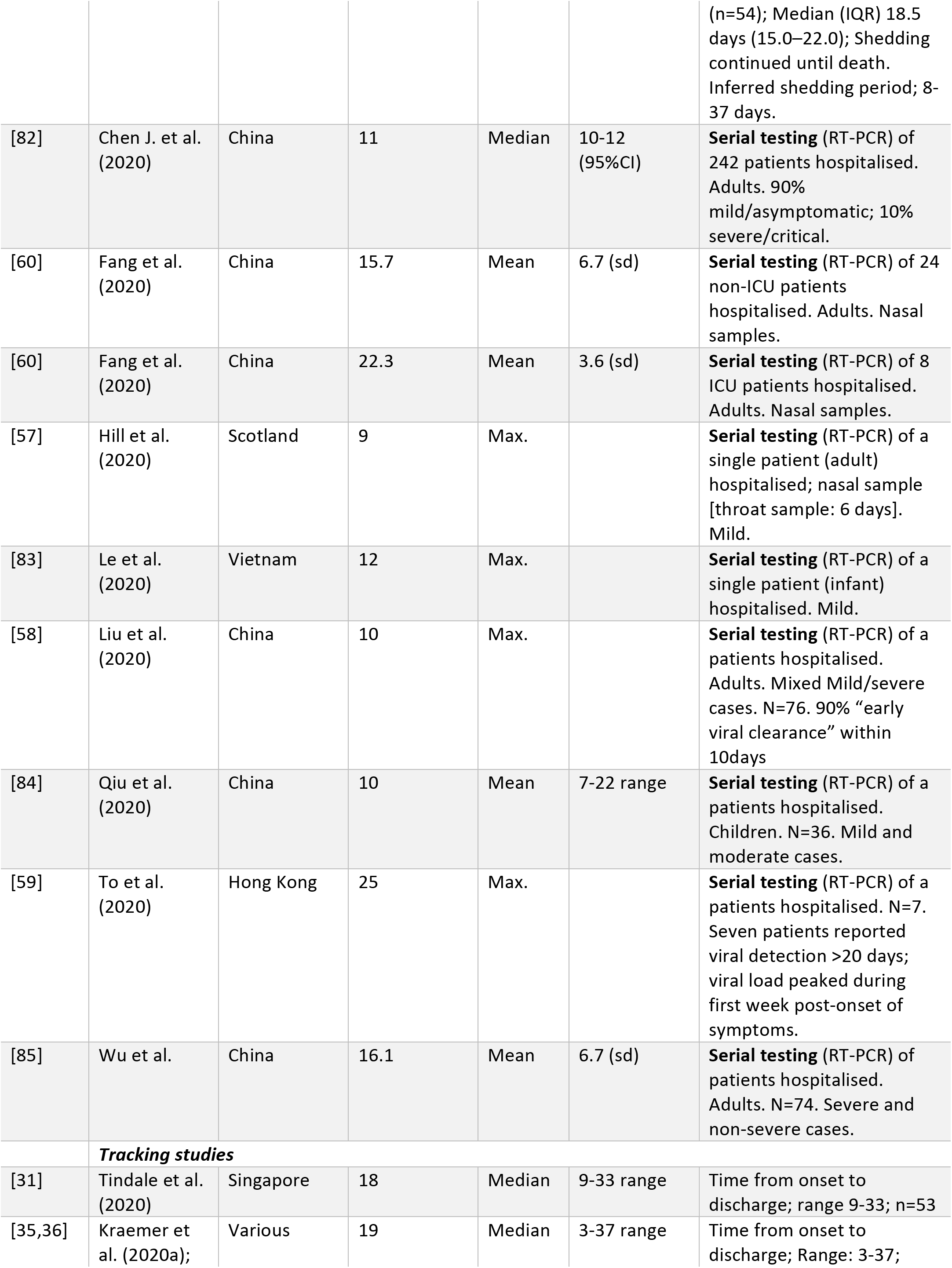
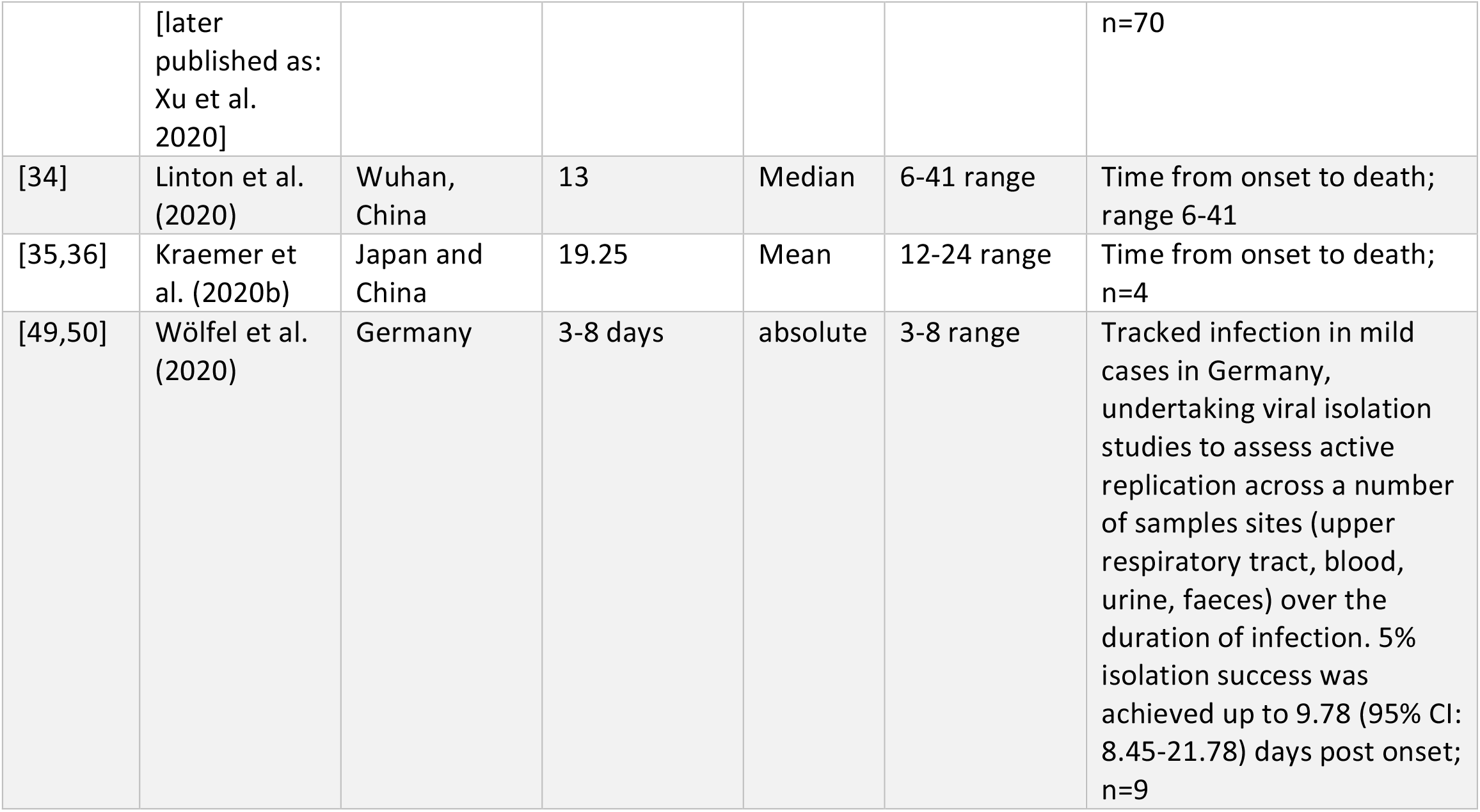
Reported infectious period (IP) for post-symptomatic cases (T5 parameter) from virological studies where serial diagnostic tests were undertaken to infer IP [onset to ≥2 tests]; tracking studies where IP is inferred from patient histories from onset to recovery or death; modelling studies where IP is reported as a prior (assumed parameter value) or an posterior estimate.

#### Infectious period for asymptomatic cases (T2)

The overall distributions and point estimates from studies for T2 are presented in Figure 1 and Table 1.

**Figure 1:**
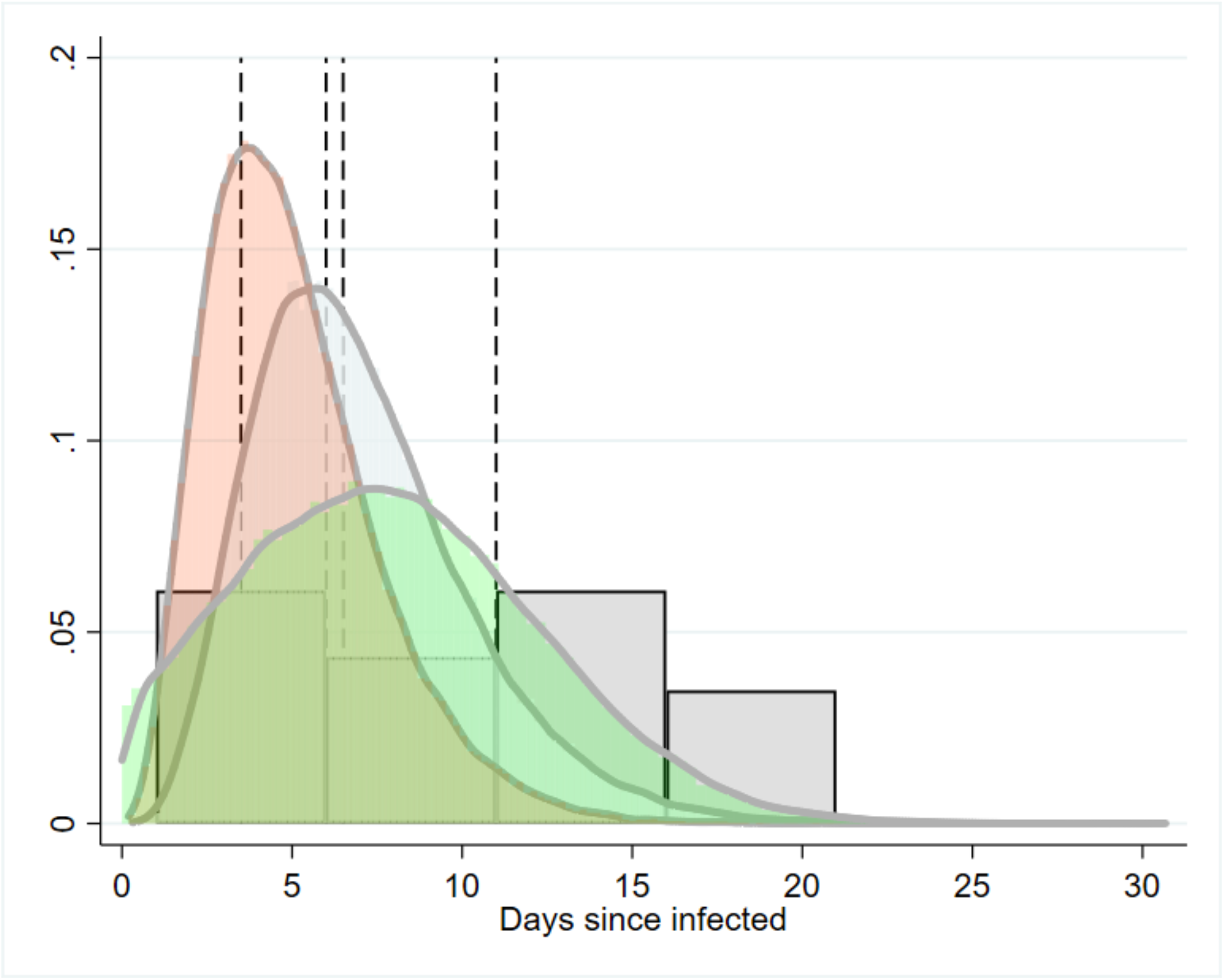
Simulation of the parameter distribution inferred for duration infectious period for asymptomatic cases (T2) inferred infectious period for Davies et al. (2020a), grey/blue curve, Davies et al. (2020b) pink curve [model priors]. Green curve: Ma et al. (2020). Histogram is the distribution of asymptomatic cases to two clear tests reported by Hu et al. (2020). Reference lines are point estimates reported from Zhou et al. (2020), Li et al. (2020), and Tuite et al. (2020a & b).[7,8,14,15,26,27,39,71]

Two virological studies reported on infectious period based on serial diagnostic testing, for asymptomatic cases, were found to have informative data. One of these studies reported on only one asymptomatic case, with exposure to negative tests being 11 days (Zhou et al, 2020). This duration should be considered an over-estimate, given that a latent period is not taken into consideration. Hu et al. [7] tracked infections of close contacts to infected persons and considered patients asymptomatic at time of diagnosis. Infectious period was defined as time from diagnosis to the first of two clear tests, providing a median duration of 9.5 days (n=24) range: 1 – 21; 3.5-13.0 IQR.

Importantly, Hu et al. [7] found that the infectious period was different between those who subsequently exhibited symptoms (i.e. pre-symptomatic) and those who did not: The median duration for asymptomatic infectious was 6.0 days (IQR: 2.0 - 12.0; N=19). This was reduced to 4.0 days (2.0 - 15.0) for cases that were asymptomatic without abnormal computed tomography (CT) scans (n=7).

Two tracing studies provide informative data (Table 1; [7,8]). Infectious period was inferred indirectly from data provided in Ma et al. [8], whereby infectious period was estimated as the difference between the upper latent period estimate minus the serial interval. Ma et al. [8] reports on 49 asymptomatic cases and inferred serial interval from infector-infectee pairs. Serial interval was calculated by assuming “onset” was at first diagnosis. Hu et al. [7] reported on a case-study cluster of infection within a house where the primary case was asymptomatic. Secondary infections occurred 4-9 days after index case exposure, the index patient tested positive until day 29 post exposure.

Modelling studies that have attempted to fit differing parameters depending on the severity of symptoms have used differing nomenclature, for example asymptomatic, “mild” or subclinical cases (Table 1).[14,15,26,27] Two papers by Davies and colleagues [14,15]model this parameter as a gamma distribution with a mean periods of 5-7 days (Fig. 2); importantly, these papers assume infectious period is the same for asymptomatic and symptomatic cases.

**Figure 2:**
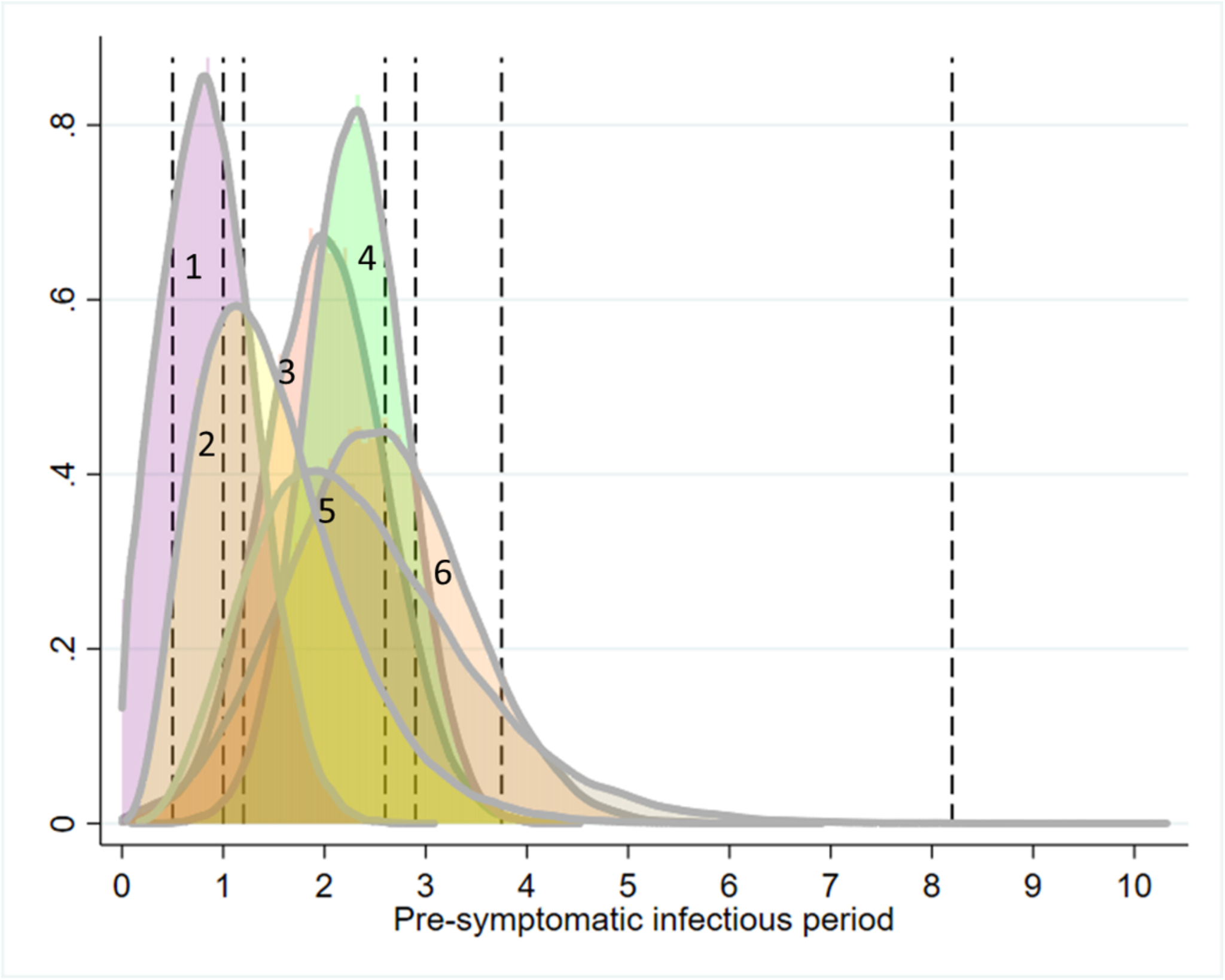
Simulation of the parameter distribution used for T3 (the duration of the pre-symptomatic infectious period for those infected individuals who subsequently develop symptoms). Curves represent simulated approximations of distributions, given information provided from primary literature. Vertical lines represent point estimates where distributions could not be inferred (see table 2). 1. Peak et al. [posterior]; 2. Davies et al. 2020b [prior]; 3. Rothe et al. 2020; 4. He et al. 2020; 5. Davies et al. 2020a [prior]; 6. Wei et al. 2020. [9,14,15,29,30,32]

#### Pre-symptomatic, infectious period (T3)

Pan et al. [3] and Hoehl et al. [28] describe the cases of two individuals tracked and serially tested by real-time reverse transcriptase polymerase chain reaction (RT-PCR) after being exposed to a patient with confirmed infection. In the latter study, the virus was isolated from samples, indicating transmission potential.

Four studies from China, Germany and Singapore provide informative data through tracing infections from cluster of infections, and through infector-infectee pairs (Table 2).[4,9,29,30] These papers included the study by Rothe et al. [9], which clarified that an asymptomatic patient visiting Germany from China may have actually experienced very mild symptoms around the time of transmission occurred (see discussion).

Five modelling papers incorporated pre-symptomatic infectious period reported as prior distributions or estimated as a model output. Two papers describe the prior distribution using a gamma distribution.[14,15] Tindale et al. [31] provide mean point estimates under four different scenarios (two populations, early and late epidemic period). Peak et al. [32] derives estimates of the pre-symptomatic infectious duration from a model of serial interval, and report scenarios where there are pre-symptomatic infectious periods.

The approximated distributions are simulated in Figure 2, which demonstrates the between-study heterogeneity in this parameter. The point estimates primarily cluster around the central tendencies of the distributions, except for Tindale et al. [31], for a model reporting for late occurring cases in Tianjin, China (8.2 days).

#### Post-symptom onset, infectious period (T5)

The T5 parameter was informed from three lines of evidence from empirically driven studies:

- time from symptoms onset to the first of two clear RT-PCR tests
- time from symptoms to hospital discharge
- time from symptoms to death

Figure 3 presents the forest plot for the mean time from symptom onset to clearance, based on serial testing meta-analysis (n=15). The mean estimated duration was 13.4 days (95%CI: 10.9-15.8). There was high heterogeneity across studies (Cochrane’s Q; p<0.001; *I*^*2*^>75%). A random effects (RE) meta-regression model suggested significant variation depending on whether studies included children as part of the sample (n=15 studies; Proportion of between-study variance explained Adj. R^2^ = 43.8%). Overall, the model estimated studies including children had on average 5.8 days shorter duration than adult only studies (95%CI: 1.7-10.0; p=0.040; SE(p)=0.003). A second univariate RE meta-regression model suggested that there was non-significant increased mean duration of 4.0 days (95%CI: −0.6-8.6; p=0.111; SE(p)=0.005; Adj. R^2^ = 22.0%; n=14) for studies that included moderate-severe or severe cases, relative to mild or mild-moderate severity cases.

**Figure 3:**
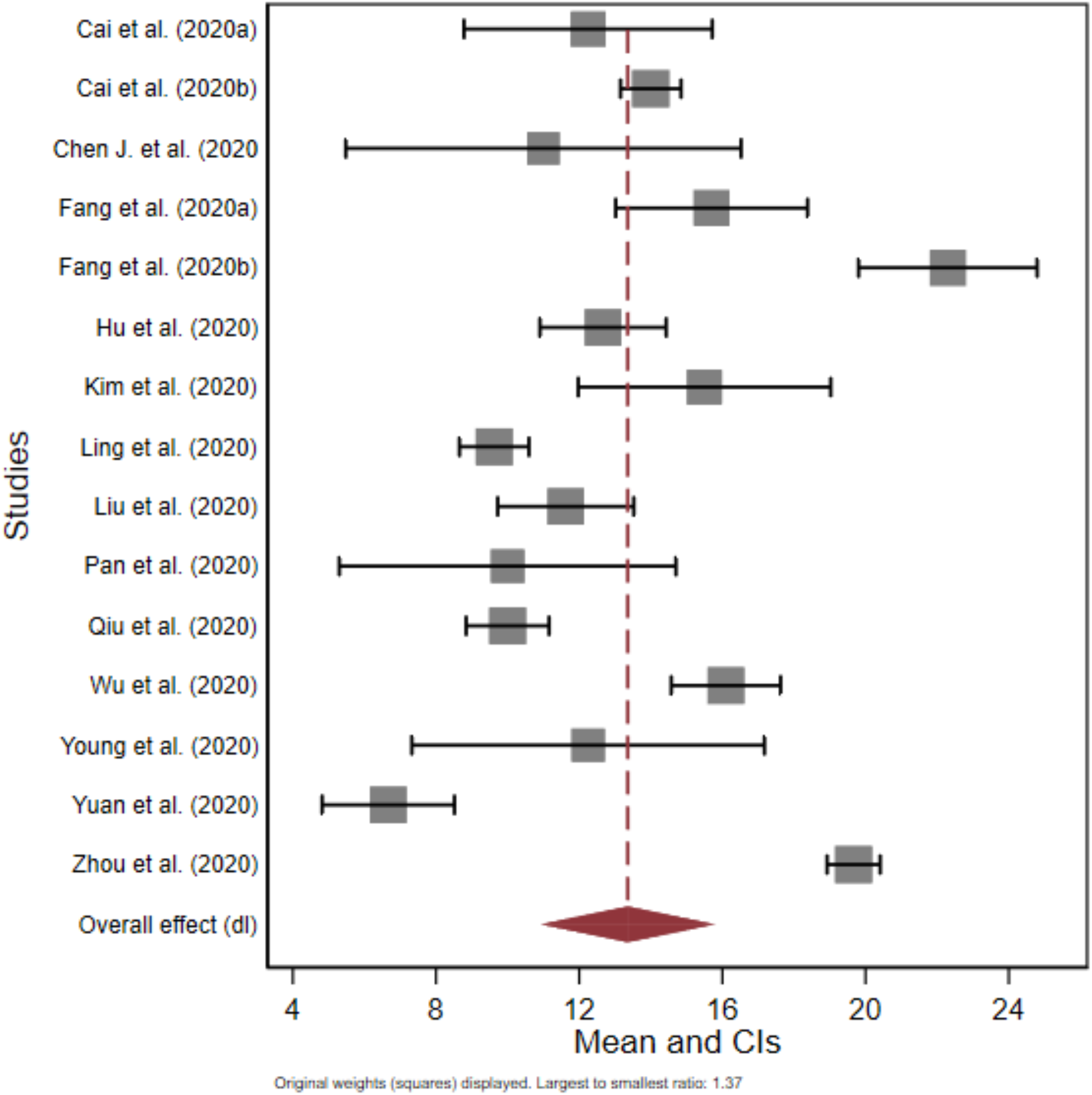
Forest plot comparison of the mean duration from onset of symptoms to death or recovery (T5) based on virological studies

High transmissibility during the first 5 days post symptom onset was described by Cheng et al. [33], based on secondary attack rates for 12 infector-infectee pairs. No contacts (n=1043) with primary cases were infected after five days of the index case onset of symptoms, inferred by the authors to suggest transmission occurring at symptom onset (but conceivably also suggest pre-symptomatic infection). Based on a cumulative density function, the authors suggest that infectiousness declines rapidly from onset of infection (distribution was truncated at 30 days); estimated cumulative infectiousness was 66.9% (95%CI: 28.7-94.8) by day 1, and reached 86.9% (95%CI: 64.3-99.5) by day 5 post-symptom onset (Figure S2).

For tracking studies relating to time to hospital discharge or death, raw case level data were available (studies n=3).[31,34–36] Histograms of the raw data are presented in Figure 4, along with the aggregated distribution. A random effect model suggested a mean duration of 18.1 days (95%ci: 15.1 – 21.0). However, there was significant variation across studies, with time to discharge being 4.96 days shorter (95%CI: 2.15-7.76; [35]), or 3.79 days shorter (95%CI: 0.8-6.7; [31]), than time-to-death [34].

**Figure 4:**
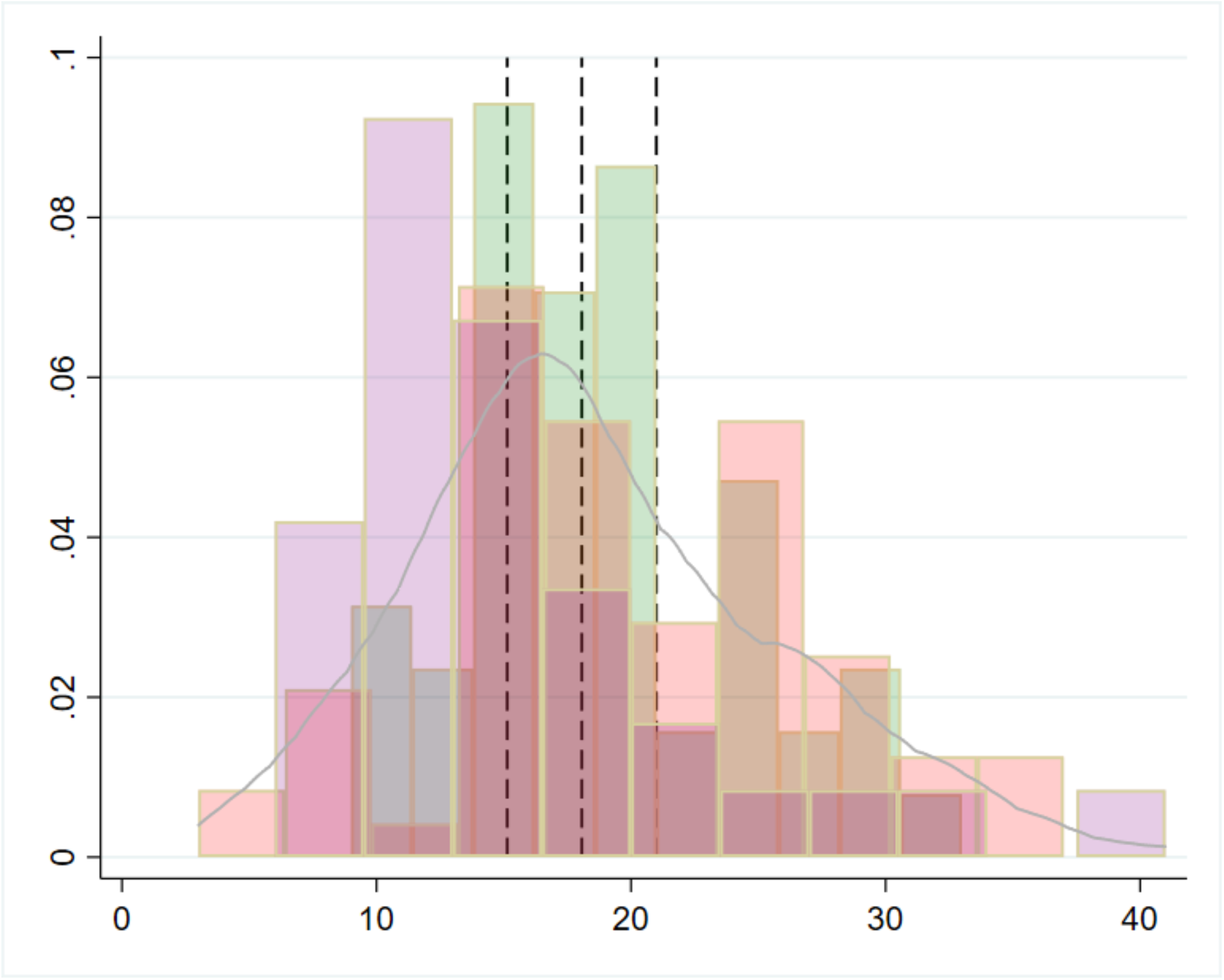
Frequency distribution of **T5**, time from onset of symptoms to recovery (here hospital discharge or death), using patient level raw data from Kraemer et al. ([35,36]; pink bars), Linton et al. ([34]; purple bars) and Tindale et al. ([31]; green bars). Blue solid line is the kernel density of the aggregated dataset Dashed lines represent the mean and 95%CI from a random effects regression model.

Two modelling papers use priors (mean: 3.2-3.5 days) to represent clinical infectious period.[14,15] However, the distribution for this parameter is right censored when patients are hospitalised or isolated and therefore not an estimate of the full infectious period *per se*.

#### Infectious period for symptomatic cases (T3+T5)

Two tracing studies supplied parameter estimates for the full infectious period for patients who develop symptoms. [8,29] He et al. [29] inferred from a publicly available dataset of 77 infector-infectee pairs that infectiousness began 2.3 days (95% CI, 0.8–3.0 days) prior to symptom onset, peaking at 0.7 days (95% CI, −0.2–2.0 days), and continued up to 7 days from onset. The authors suggest that the transmission risk diminishes 7 days post symptom onset. This suggests that the average infectious period, assuming a symptomatic infectious period of 7 days was approximately 9.3 days (7.8-10 days 95%CI, where CI is only reported for the pre-symptomatic period). He et al. [29] estimated that the proportion of all transmission that was pre-symptomatic was 44% (95% CI, 25–69%). Ma et al. [8] analysed data from a number of countries (China, Germany, Japan, Malaysia, Singapore, Vietnam), collating 1155 cases from public data. They estimate several parameters, including “maximum latent period” and the serial interval. The authors estimated the infectious period as maximum latent period minus the serial interval. Given their parameter estimates and methodological approach, infectious period would have been 5 days (range 0-24; IQR: 2-9; calculated from data presented within the paper).

Seven modelling papers reported duration of infectious period (T3+T5; Table 4), with the reported central tendency for the distribution varying from 3-20 days. The form of the distribution offered to models for this parameter varied considerably, including point estimates (deterministic models), flat (uniform), Gaussian, Weibull and gamma distributions. Li et al. [27] estimated the shortest median duration of 3.45 days, with a flat (uninformative) prior distribution corralled between 3-5 days. In contrast, Zhu et al. [37] used a mean prior of 10 days, with the model estimated mean duration being 12.5 days (variance 10; Weibull distribution). Piccolomini and Zama [38] used a fixed estimate of 20 days infectious period, to model the Italian epidemic. Two papers from the same group [14,15] suggested that infectious period for asymptomatic cases approximated for symptomatic cases where there was no right censoring (that is, transmission being halted through isolation or hospitalisation; gamma distributions of mean 5 or 7 days). Tuite et al. [26,39] also assumed the same duration for “mild” and “severe” symptomatic cases (6-6.5 days).

**Table 4:** Reported infectious period (IP) for symptomatic cases (T3+T5 parameter) from virological studies where serial diagnostic tests were undertaken to infer IP [exposure to ≥2 tests]; tracking studies where IP is inferred from patient histories from onset to recovery or death; modelling studies where IP is reported as a prior (assumed parameter value) or an posterior estimate.

| No. | Study | Location | Parameter (days) | Central tendency reported | Variation (days; inclus.) | Comment |
| --- | --- | --- | --- | --- | --- | --- |
| **Tracking studies** |  |  |  |  |  |  |
| [29] | He et al. (2020) | Vietnam, Malaysia, Japan, China, Taiwan, USA, Singapore | 9.3 days | Mean | 7.8-10 (95%CI*) | The paper reported on 77 infector-infectee pairs which were sequential/serially tested, using publicly available data. Viral dynamics (Guangzhou, China; N=94) interpreted by the authors suggested an infectious period starting 2.3 (95% CI, 0.8–3.0 days) days prior to symptoms, peaking 0.7 days (95% CI, –0.2–2.0 days), continuing up to 7 days from onset. * CI from pre-symptom infectious period only. |
| [8] | Ma et al. (2020) | Various | ~5 days | Median | Range 0-24 | The authors estimated the infectious period as latent minus the serial interval, using a dataset of 1155 cases. Range 0-24; IQR: 2-9; calculated from data presented within the paper. |
| **Modelling studies** |  |  |  |  |  |  |
| [27] | Li et al. (2020) | China | 3.45 days [posterior estimated from model for documented cases] | median | 95%CI for the mean: 3.19, 3.72 | Mathematical model. Priors for <u>mean</u> documented infectious period was a flat [uniform] distribution 2-5. ‘Documented’ cases were defined as those severe enough to be confirmed. This corraling of the infectious period relative to other studies should take into account that the distribution is used for the central tendency, not the whole distribution. |
| [26,39] | Tuite et al. (a, b) (2020) | Canada | 6-6.5 days [prior; fixed parameter within a deterministic model] | Fixed parameter |  | Mathematical model [deterministic], with a fixed parameter estimate of 6.5 days (a) and 6 days (b), respectively. Important to note that duration for 'mild' was equal to severe cases. |
| [86] | Lourenco et al. (2020) | UK | ~3-5 days [posterior; approximate depending on scenario tested] | mean | 95%ci of 3-6 days | Mathematical model. The <u>prior</u> used was given a Gaussian distribution (normal curve); mean 4.5; SD 1; approximate 95%ci of 3-6 days. The reported posterior of this parameter was presented graphically and depended on R0 and proportion at risk. Depending on the scenarios tested, mean duration of infectiousness appeared to vary from 3-5 days. |
| [37] | Zhu et al. (2020) | Wuhan, China | 12.5 days [posterior estimated from model] | Mean | 11.4 variance | Mathematical model. The parameter was estimated using a Weibull distribution. The prior for this parameter was 10 days. The posterior variance around the mean was 11.4, and therefore the distribution had a long tail. This study was a modelling [SEIR extended model]. |
| [15] | Davies et al. (b) (2020) | UK | 7 days [Prior] | Mean |  | Model with asymptomatic infection compartment. Modelled with a gamma distribution, beta 1.4; alpha 5. Despite, the subclinical aspect of this parameter, it could be considered analogous to total infectious period without intervention. |
| [14] | Davies et al. (b) (2020) | UK | 5 days [Prior] | Mean |  | Model with asymptomatic infection compartment. Modelled with a gamma distribution, k=4. Authors: "Assumed to be the same duration as total infectious period for clinical cases, including preclinical transmission" |
| [38] | Piccolomini and Zama (2020) | Italy | 20 days [Prior] | Fixed |  | Parameter estimate assumed for the infectious period within an SEIRD model, fitted to data from the epidemic in Italy. |

### Viral load dynamics

Viral load was reported from 21 papers using real-time reverse transcriptase polymerase chain reaction (rRT-PCR) testing, generally post-symptomatic monitoring.[3,29,40–59] Qualitatively, the viral dynamics described early increase in viral load, peaking around onset or within 2-4 days of symptom onset (Figure 6 for a theoretical model), before decreasing gradually over the next one to three weeks post symptom onset. Maximum duration of detection ranged from approximately 20-49 days, with the longest duration associated with faecal samples (see below for discussion). The duration where ribonucleic acid (RNA) was recoverable by RT-PCR may have been truncated due to insufficient follow-up in some cases. Studies that have investigated blood samples have provided some evidence for an association with severity of infection [16,60], though it is not clear whether this is a consistent feature of SARS-CoV-2 infection [40].

It should be noted the lack of data on pre-symptomatic or asymptomatic cases with regards viral load. An exception was Kam et al. [61] who describe a pre-symptomatic case in an infant. In another study, Zou et al. [53] undertook serial RT-PCR testing from nasal and throat swab samples from 14 imported cases, and 4 secondary cases, in Guangdong, China. The dynamics of the infection in terms of cycle threshold (Ct) values and RNA copy number were described; Ct values of 30.76, 27.67, 24.56, and 21.48 corresponding to 1.5×10^4^, 1.5×10^5^, 1.5×10^6^, and 1.5×10^7^ copies per milliliter. Hence, lower Ct values infer higher viral loads. The authors report on a patient without symptoms, but with positive nasal swabs (Ct values, 22 to 28) and throat swabs (Ct values, 30 to 32) testing positive on days 7, 10, and 11 after contact. Importantly, the authors suggest “the viral load that was detected in the asymptomatic patient was similar to that in the symptomatic patients.” Furthermore, Kimbell et al. [62] report that Ct values between asymptomatic (21.9 to 31.0), pre-symptomatic (15.3 to 37.9), and symptomatic cases (18.6 to 29.2) within a nursing home environment did not differ significantly. To et al. [59] present data on temporal profile of viral load from saliva samples, and found that median initial and peak viral loads in severe cases were non-significantly higher (p>0.5) by approximately 1 log10 higher than those in mild cases. Liu et al. [58] present data showing viral load being 60 times greater for severe cases relative to mild cases.

This lack of pre-symptomatic data may result in left truncation of the risk distribution associated with viral load and shedding. Therefore, the typical timing of peak viral shedding (whether prior to, at, or after onset), and it’s impact on transmission, is still uncertain. He et al. [29] reported highest viral load at symptom onset from patients sampled in a hospital in China. Furthermore, the author’s estimate using a separate infector-infectee dataset (n=77) that 44% (95% CI: 25–69%) of infectee cases were infected during the pre-symptomatic stage of the infector. Separately, a modelling paper by Ferretti et al. [63] also appears to support this, estimating that 47% (0.9/2) of total transmission contributing R_0_, an overall measure of transmission during an infection, was pre-symptomatic (also see [33]).

Wölfel et al. [50] provides important data on a cohort of nine ‘mild’ cases which were serially tested using sputum, swabs (throat and nasopharyngeal), urine and faecal samples over time. Importantly, the virus was isolated, and inferences on viral replication could be made. Viral Isolation, and insights into viral replication, improve inference around viral dynamics and transmission risk. The study suggested high viral loads shortly after symptom onset, which declined thereafter over time. Positive cultures were found from day 3-8 post-symptom onset (Figure 5), and the minimum 5% isolation success was achieved up to 9.8 (95% CI: 8.5-21.8) days post onset from throat and lung samples but not faeces, blood or urine.

**Figure 5:**
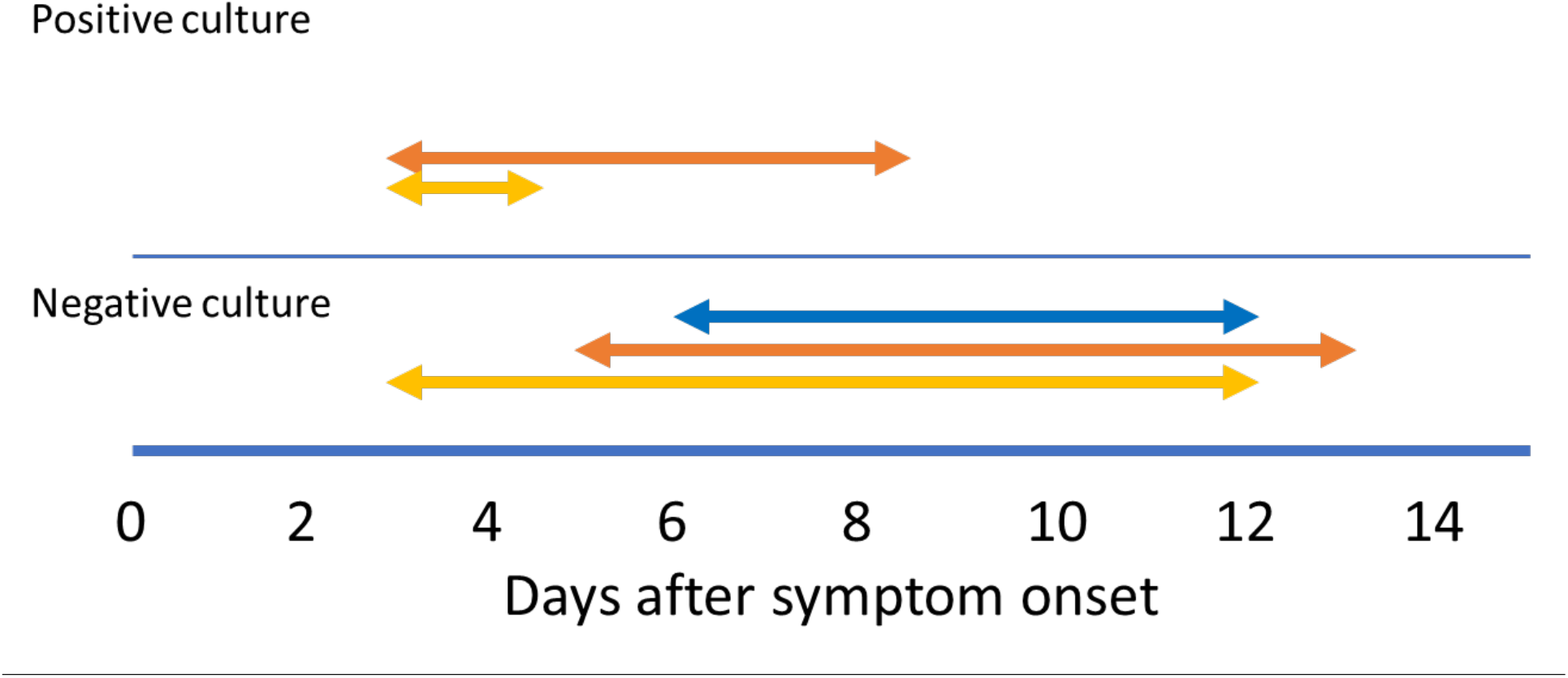
Timeline for positive culture results of SARS-COV2 from throat, sputum and stool samples; Yellow line = Throat swabs; Orange line = Sputum samples; Blue line = Stool samples; Adapted from Wölfel et al.[50].

## Discussion

Inferring infectiousness was challenging given the heterogeneity of evidence available. Virological diagnostic studies provide robust time series of infection, however, is limited by inferring the relationship between PCR diagnostics and infectiousness. These data can also be affected by sampling procedure and sample sites (e.g. upper respiratory, lower respiratory, faeces, urine, blood). We have excluded RT-PCR durations based on faecal sampling due to the uncertainty whether these data pertain to transmission potential ([50]; see below). Virological studies where culturing has taken place, and where viral replication can be inferred would also be considered superior data to infer infectious period, relative to estimates of viral load alone.[50] Where this has taken place, the data would suggest average infectious periods of up to 9.8 days post-symptoms. Recent modelling work suggest that the duration of viral detectability could overestimate the infectious period somewhere between 2-6 days.[64]

Viral load studies suggest peak viral load occurs close to symptom onset (potentially, −1 to 7 days of onset), however there is uncertainty whether this typically occurs prior to, on, or after onset (Figure 6 for conceptual model). High viral loads, measured as Ct values, have been recorded for one week to 20 days post symptom onset, with a general decreasing trend with time. For example, To et al. [59] estimates a declining slope per day for log10 RNA copies per ml of −0·15 (95% CI −0·19 to −0·11; *R*^2^=0·71). There are some studies reporting associations between viral load and symptom severity, with higher metrics of viral load in severe cases.[3,58,59] However, Zou et al. [53], and more recent data from Italy,[64,65] suggest similar viral loads in symptomatic and asymptomatic cases.

**Figure 6:**
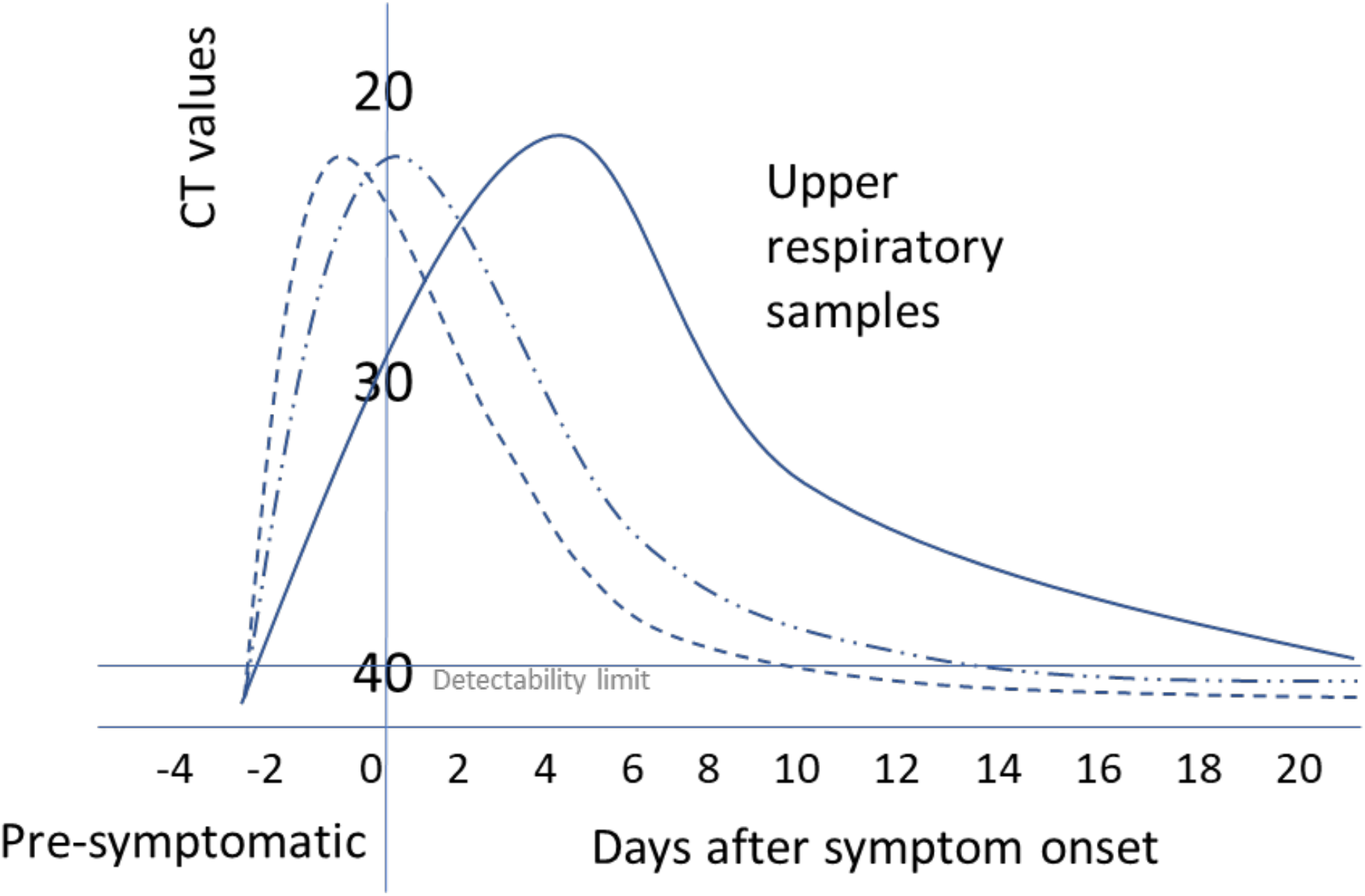
Composite inferred model for cycle threshold (CT) value changes from serial RT-PCR testing for SARS-COV2; currently uncertain whether peak viral load typically occurs prior to, on, or post-symptom onset (primary literature informing this model includes [29,50,53,59]).

We tested the hypothesis that severity of symptoms had an effect on symptomatic infectious duration using a meta-regression approach. There was a trend towards studies that included severe cases tended to have longer duration (estimated to be 4.0 days longer), but the effect was not significant. Some studies have reported an association between duration of infectiousness and severity (e.g. [58]). But uncertainty of whether this is robust remains.

Virological studies that included children (either mixed adult children, or children only cohorts) appeared to have shorter T5 durations (estimate: 5.8 days shorter). Liao et al. [66] present data which suggests that children and ‘young adults’ (<35 years old) infected cases exhibited long incubation time (exposure to symptom on-set; mean 7.2 days), and short serial interval (mean 6.5 days; median 1.9 days; time from onset in primary to onset in secondary case).

Contact tracing studies provided robust evidence of transmission events, and therefore infectiousness, but can be limited by the inferred timing of events, and symptoms experienced, due to the self-reported nature of data collection (recall bias). The subjective nature of self-reporting indeed can have an impact on case definitions of ‘asymptomatic’, which has led to some doubt on asymptomatic transmission in one case.[9] Rothe et al. [9]describe a case of apparent asymptomatic transmission from a Chinese visitor to business associates in Germany, which was cast into doubt when health officials reported that the patient had indeed experienced some, albeit minor, symptoms.[67] Rothe et al. [9] subsequently updated the clarification of the patients self-reported symptoms during the presumed asymptomatic infectious period, which included “feeling warm” and “feeling cold”. However, the patient only “recognized getting sick” after she returned to China on day four after the presumed exposure event.

Modelling parameters provide information on how COVID-19 data are being used and interpreted in the research community, given the limited data available. Posterior estimates also provide information on the parameter space at which infectious period central tendency reside, given other parameters and assumptions in the model. Models used highly varied approaches to modelling infectious period, which in turn resulted in highly variable parameter estimates used to inform the studies.

### Overall duration findings

There are few data for the precise definition of the asymptomatic infectious period (T2) parameter. Some reported asymptomatic cases can actually be pre-symptomatic, when cases are subject to follow-up (e.g.[66]; see discussion above). However, Hu et al. [7] do provide the data for asymptomatic cases [that remain asymptomatic] across their presumed infectious period. Therefore, in the first instance a parameter mimicking their data is probably the best available data. Note, there is a large variation in this data parameter, and a gamma distribution of a shape alpha 3, beta 2, mean 6, may be appropriate for the initial model runs. Despite these being the primary informative data, caution is required, given the uncertainty around the relationship between RT-PCR results and infectiousness. Overall, an informed central tendency of ∼6 days, with very low probability draws for durations >20 days for the T2 parameter may be considered given the current state of knowledge.

The pre-symptomatic period is sometimes referred to as ‘preclinical infectious’ period (parameter T3). This has been estimated from several papers, and the central tendency of these estimates vary from <1 - 4 days, cautiously approximating to 2 days, on average. The maximal reported period for T3 from any population, was reported by Tindale et al. [31] at 8.2 days. Current models have used central tendency estimates of 0.5 to 2.4 days.[14,15,26,39] It should be noted, that this period could also be measured as the difference between incubation and latent period, or the difference between serial interval and incubation period.[12] The relative consistency around the duration of this period allows for some confidence of its distribution. Current understanding of viral dynamics of infection suggest that viral load and shedding increases during post-latent phase, peaking around onset [for symptomatic cases], before declining.[29,50,53] This aspect of the natural history of infection may be important when attempting to model transmission dynamics.

Length of infectious period in symptomatic cases that do not isolate (T5 parameter) has also been rarely directly measured in the literature, as serial monitoring of patients in terms of symptoms or viral load (rt-PCR) generally occurs after diagnosis and/or after admission to hospital [from a modelling perspective, this means cases are censored as they are assumed to no longer contribute to transmission]. If natural progression of infection after diagnosis or hospital admission mimics the course of infection for those who do not isolate, the review of the literature describing time to two clear tests is informative. Symptom onset to serial testing clearance [assessed the time to first of two RT-PCR clear tests] averaged 13.4 days (95%CI: 10.9-15.8) from our meta-analysis. In the maximal case, where patients succumb or fully recover from infection, time from symptoms to death or discharge may be informative. Studies that collated such information suggest mean durations of 18.07 days (95%ci: 15.14 - 20.99), but with time to discharge being 4.96 days shorter (95%CI: 2.15-7.76) on average than time to death. These values may represent an over estimation of the infectious period; one study suggested that there was on average 2.5 days between end of infectiousness and ‘removal’ (recovery or death).[37]

Cheng et al. [33] provided evidence of transmissibility, based on attack rate from primary to secondary cases, at around symptom onset. The authors estimate cumulative infectiousness from onset, which suggests that 67% of total infectiousness potential occurs by the first day post-onset. Most of the total infectiousness occurs within 5 days (86.9%) post onset, with the remaining infectiousness potential (13.1%) being distributed up to day 30 (this truncation is an assumption by the authors). It is possible that pre-symptomatic transmission occurred during this study, but the authors do not estimate what proportion of transmissions occurred during a pre-symptomatic infectious period, or its potential duration.

A model by He et al. [29] is informative for overall symptomatic duration (T3+T5), using 77 infector-infectee pairs where COVID-19 transmission occurred in China. The study reported that infectiousness was apparent on average 2.5 days prior to symptoms, reached a peak in risk at 0.6 days before symptoms, and decline up until 7 days after onset (9.5 days total infectious period). The proportion of transmission before symptom onset (area under the curve) was estimated as 44% (95% CI, 25–69%), based on inferences on incubation period. The authors suggest their data supported the view that transmission risk decline substantially after 7 days post-symptoms onset.

Model estimates used for infectious period parameter appears to be shorter than virological studies tracking RNA viral load over time. For example, Liu et al.[27] fitted a flat prior distribution for mean duration (*D*) fixed to vary between: 2 ≤ *D* ≤5 days, and Lavezzo et al. [64] fixed infectious period to 2 days in their epidemic model; whereas viral repeat testing studies provide evidence to suggest high viral loads can be detected to up 20 days [e.g. pharyngeal swabs], and potentially longer from faecal samples (up to 3-4 weeks post symptoms onset). Oral-faecal transmission risk is currently unknown, but some doubt has been raised about studies that have reported positive RTPCR test results (see [68]; but there may be some evidence of the risk amongst children; [69]). Wölfel et al. [50] has produced an important study that provides some data on viral replication, and the site and duration over which this may be taking place. Their data suggests that viral replication, with high viral loads, occur in the upper respiratory tract, over the first week of symptoms peaking in day 4. Virus could not be isolated from faecal samples, despite high RNA concentration. Furthermore, virus was not isolated from blood or urine in that study.[50]

### Study limitations

Overall, the studies included were of good quality, though due to the rapid need for information from the global research community many papers are pre-prints that have yet to be reviewed (at time of writing). Many papers were limited in terms of sample sizes, with several papers being case studies of one patient or single cluster outbreaks. There was a diversity of methods employed to infer dynamics of infectiousness across studies, and therefore the evidential base was variable. Some issues around nomenclature were noted, including definitions of asymptomatic, infectious period, latent, and incubation period. It is possible the same data may have been used across different studies, especially where publicly available data were used.

There was significant heterogeneity across study findings, and this was related to diversity of clinical findings and methods employed. The meta-analysis employed for one parameter (T5) using virological studies, where cross study comparisons could be made, suggested that the heterogeneity was high. Fu et al.[70] cautions against combining studies to give an overall estimate without exploring subgroup or meta-regression analysis, which we have done here. The meta-regression was based on a small number of studies (n=12-13). Cochrane’s handbook suggests 10 studies for each level of a meta-regression, however in practice much lower numbers have been used to test hypotheses [22], as is the case here. Fu et al. [70] recommend a minimum of 4 studies per category, and therefore we dichotomised our predictor variables to ensure we met this minimum. Aggregating our categories resulted in crude findings.

Another limitation is that a systematic review was not undertaken to inform this research, hence there is a possibility that some relevant studies were overlooked. However, comprehensive search strategies were conducted by two independent research groups to inform this research, hence limiting the potential for missing key studies.

## Conclusion

There are few data to inform asymptomatic infectious period (T2 parameter). One study provide data that suggest a median period of 4-9.5 days, however, given the viral dynamics, this distribution could have an extended tail with low probability long infectious periods of up to 20 days. The pre-symptomatic infectious phase (T3) is quite narrowly defined to a mean of approximately 2 days (range: <1-4) within the literature. However, there is great uncertainty around the infectious period from onset to recovery or death (T5 parameter). The symptom onset until clearance (based on two negative RT-PCR tests) parameter estimate of 13.4 days (95%CI: 10.9-15.8) is informative for T5 parameter, only if one assumes that RT-PCR positive results equate to having infectious potential. Many current models corral the infectious period to shorter time periods than what virological studies have suggested, with one recent study suggesting that duration of viral detectability over-estimates the infectious period on average by 2-6 days. While viral RNA can be detected for long periods of time, especially from faecal samples, the ability to isolate the virus ifrom nfected cases quickly declines after one-week post-symptoms. Some modelling papers have assumed that infectious period is invariant to whether cases are asymptomatic or symptomatic, however, the data available are not yet rich enough to inform whether this is a good assumption. Similarly, it is not yet established whether viral loads are similar between asymptomatic and mild, moderate, or severe symptomatic cases, with conflicting reports in the literature.

## Data Availability

Data are available within the paper or supplementary material

## Author contributions

AWB conducted the eligibility screening of shortlisted studies, extracted the data and conducted the analyses with input from all authors; ÁC, KH and FB conducted the initial literature searches; DM, KOB, KW conducted searches and screened shortlisted studies; AWB completed the initial draft of the manuscript; CM reviewed the statistical methods; CM and MC undertook quality control interim review; All authors read and approved the final manuscript.

## Data statement

The data and code are presented in supplementary material

## Patient and public involvement statement

It was not appropriate or possible to involve patients or the public in the design, or conduct, or reporting, or dissemination plans of our research

## Supplementary material

**Figure S1:**
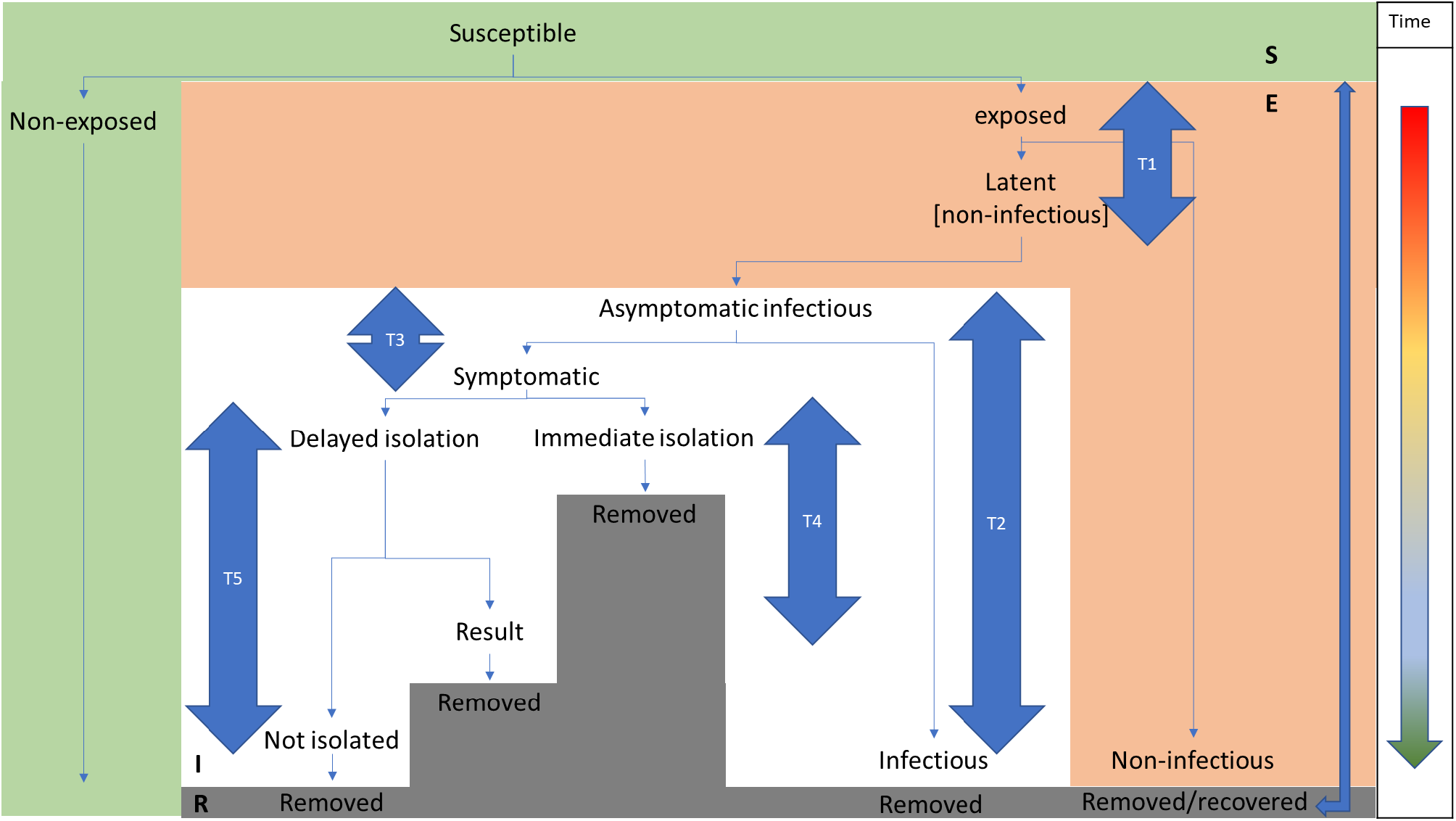
Conceptual model of the key temporal parameters impacting COVID-19 infection progression over time. T1: Latent period; T2: Asymptomatic infectious period; T3: Pre-symptomatic infectious period; T4: Symptom onset to diagnosis [self-isolation] or hospitalisation; T5: Symptom onset to removed [death or recovery]

**Figure S2:**
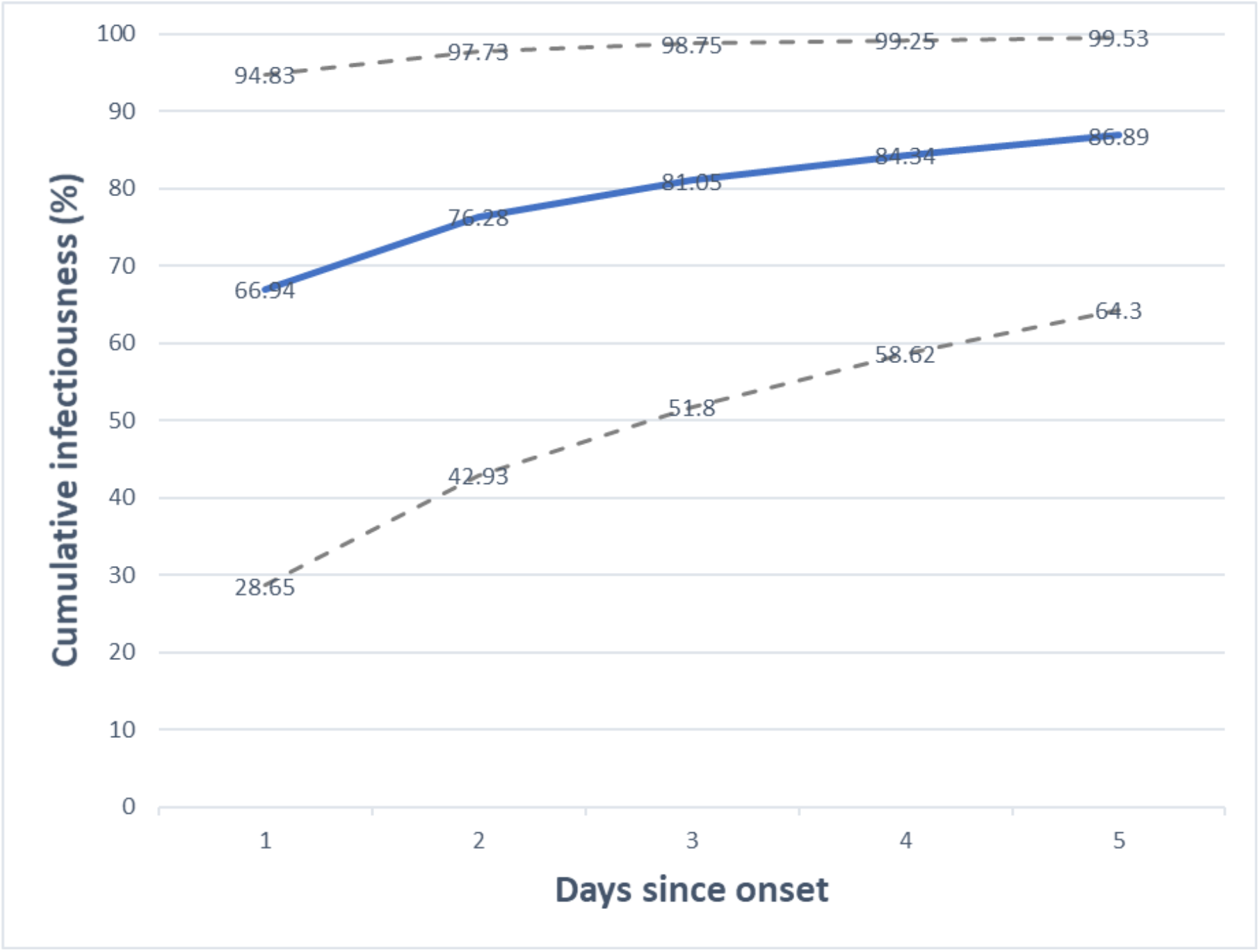
Cumulative infectiousness (% of total infectiousness) based on infector-infectee pair data in the paper by Cheng et al. 2020. The accumulation curve is based on a gamma density function, coupled with a probability function to capture the maximal probability if exposed to a primary case.

